# Unipolar voltage for better characterizing left atrium substrates: Comparing the predictive efficacy for recurrence post atrial fibrillation ablation in a post-hoc analysis of STABLE-SR-III

**DOI:** 10.1101/2024.02.07.24302471

**Authors:** Xiuyu Qi, Hongwu Chen, Gang Yang, Mingfang Li, Kai Gu, Hailei Liu, Zidun Wang, Xiaohong Jiang, Chang Cui, Cheng Cai, Minglong Chen, Weizhu Ju

## Abstract

**Background:** Intracardiac mapping has become a prevalent technique for assessing cardiac fibrosis. While bipolar recording is universally acknowledged as an indicator of cardiomyocyte activation, unipolar recording has emerged as an alternative technique due to its advantage of providing a wider field of view. This study aims to compare the efficacy of unipolar voltage (UV) versus bipolar voltage (BV) in predicting recurrence in elderly atrial fibrillation patients.

**Methods:** In Substrate Ablation in the Left Atrium during Sinus Rhythm Trial III, 414 patients were enrolled in the modified intention-to-treat analysis. Of them, 375 patients who completed the follow-up with preserved mapping data were included in the analysis. For each patient, the mean UV and BV was obtained from the electrograms sampled in left atrium (LA).

**Results:** Both low UV and BV of LA had significant associations with the long-term recurrence of atrial tachyarrhythmia (ATa). At the same time, only mean UV was independently associated with the outcome. The model by UV with ablation feature had higher discriminatory power to predict ATa recurrence compared with BV model (AUC: 0.858 vs 0.757, P<0.001). Decision curve analysis demonstrates that UV model provides larger net benefit across the range of reasonable threshold probabilities between 0% and 70% compared with BV model between 0% and 45%. In subgroup analysis, UV reveals more powerful predictive efficacy compared with BV, with the AUC 0.843 vs. 0.751 (P=0.0008) in CPVI alone cohort and 0.882 vs. 0.750 (P=0.0004) in CPVI plus cohort, respectively.

**Conclusion:** UV exhibits a higher efficacy for predicting long-term ATa recurrence after ablation compared with BV in elderly patients with atrial fibrillation. The superiority exists regardless of whether the patient accepts substrate modification. The outcome suggests that unipolar recording may better characterize LA fibrosis by capturing more comprehensive transmural features than bipolar signals.

**Clinical Trial Registration:** ClinicalTrials.gov; URL: https://www.clinicaltrials.gov. Unique Identifier: NCT03462628

**Clinical perspective:** *What’s known:*

- Atrial fibrosis represents a central pathophysiological feature and has been correlated with complications and resistance to drug and ablation therapy for atrial fibrillation. Evaluating the degree of fibrosis holds paramount clinical importance.
- Contact intracardiac mapping stands out as a common method for assessing fibrosis. The amplitude of bipolar electrogram signifies the activation of viable cardiomyocytes. Moreover, the decline in amplitude of bipolar voltage has been confirmed to be associated with the long-term recurrence after ablation.

*What the study adds:*

- In comparison to bipolar voltage, endocardial mean unipolar voltage of left atrium exhibits a higher efficacy for predicting recurrence after ablation in elderly patients with atrial fibrillation.
- The superiority predictive ability of unipolar mapping suggests its advantage of providing a broader, more penetrative field of view, enabling the identification of arrhythmogenic substrates in deeper layers of the atrium.

## Introduction

Atrial fibrillation (AF) is one of the most prevalent arrhythmias^1^. Atrial fibrosis has been established as the central pathophysiological feature and has been associated with complications and resistance to drug and ablation therapy for AF^2–4^. Consequently, evaluating the degree of fibrosis holds significant importance for therapeutic decision-making and prognosis assessment^5^. Contact intracardiac mapping using electrodes has emerged as a clinically accessible modality for fibrosis evaluation. The amplitude of the bipolar electrogram indicates the activation of viable cardiomyocytes. In clinical practice, low voltage area (LVA) identified through endocardial bipolar mapping is generally regarded as indicative of atrial fibrotic regions^5–7^. Despite this, several publications had raised concerns about the “field-of-view” of bipolar electrograms, highlighting the inherent limitation of being unable to detect transmural substrates^8^.

Unipolar voltage (UV) mapping provides a distinct perspective and potential^9–11^. With its broader, more penetrative field of view, UV mapping can identify arrhythmogenic substrates in deeper layers of the heart^8^. From this point of view, we hypothesized that atrial UV may reflect more comprehensive transmural information regarding atrial substrates in patients with AF. Consequently, it could potentially offer a more robust prediction for the recurrence of patients who undergo ablation. In this post-hoc analysis of Substrate Ablation in the Left Atrium during Sinus Rhythm Trial III (STABLE-SR III)^7^, we examined the predictive efficacy of UV versus BV for the long-term control of atrial arrythmia (ATa) in elder patients with paroxysmal AF.

## Methods

### Study design and population

The study design of STABLE-SR-III (Substrate Ablation in the Left Atrium during Sinus Rhythm Trial III; ClinicalTrials.gov; URL: https://www.clinicaltrials.gov. Unique Identifier: NCT03462628) trial, have been published previously. Between April 1, 2018 and August 3, 2020, 65 to 80-year-old patients with paroxysmal AF were recruited for catheter ablation in 14 centers across mainland China. The inclusion and exclusion criteria adhere to the main study. A straightforward computerized randomization method is employed to assign patients to either the circumferential pulmonary vein isolation (CPVI) alone or CPVI plus low-voltage-area ablation arm in a 1:1 ratio. In the study group, additional ablation procedures were carried out beyond CPVI. These procedures encompassed the homogenization of LVAs, defragmentation in the transitional zones, and, if required, de-channeling was performed. In the CPVI alone group, a comprehensive voltage mapping was meticulously conducted during sinus rhythm without any supplementary ablation procedures.

The STABLE-SR-III trial was approved by the Institutional Review Broad of the First Affiliated Hospital of Nanjing Medical University and each participating site. A written informed consent was obtained from all patients.

### Sampling of the points and evaluation of the voltage

After CPVI, by using Thermocool SmartTouch and CARTO3 System (Biosense Webster, Inc., Diamond Bar, CA, USA), a detailed left atrium (LA) voltage map was created point by point under sinus rhythm. To ensure the accuracy of mapping, a contact force with over 5 g was mandatory, and at least 150 surface points should be sampled.

The BV and UV values were automatically determined and provided by the CARTO 3 system. All voltage values for all mapped points were collected offline and exported from the system. The mean voltage was calculated based on all values of the mapped points.

### Study endpoints

The primary endpoint of this study aimed to assess the freedom of any ATa without the need for antiarrhythmic drugs following a single-ablation procedure. Recurrence was specifically defined as an episode of ATa occurring beyond the 90-day blanking period and lasting longer than 30 seconds. The occurrence of ATa in the first three months after the index ablation (the blanking period) was not counted. The episodes of ATa were confirmed through a blinded review by two senior electrophysiologists.

### Statistical analysis

The long-term clinical outcomes (up to 23 months) were compared between BV-based model and UV-based model. Continuous data were presented as mean ± standard deviation. Group comparisons were performed using an independent sample t-test or Mann–Whitney U nonparametric test. Classification data were expressed as frequency or rate (%), and comparison between groups was performed by Pearson χ2 test method. The logistic regression test was performed using AF recurrence as the dependent variable. The receiver operating characteristic curve (ROC) and area under curve (AUC) was calculated to compare the discriminatory capability among the models and quantify the discrimination ability. Statistical differences in the AUCs were compared using the Delong test. The clinical utility of the model was evaluated by decision curve analysis. The calibration curve was plotted using a bootstrap resampling method (1000 bootstrap resamples), and the Hosmer and Lemeshow test was performed. A two-sided p value less than 0.05 was considered statistically significant. Data processing and analysis were performed using R version 4.3.0(http://www.R-project.org/) and SPSS, version 25.

## Results

### Baseline characteristics

438 patients were enrolled between April 1, 2018, and August 3, 2020, and were randomly assigned to receive CPVI plus LVA ablation (n=219) or CPVI alone (n=219). Among them,

414 patients were enrolled in the modified intention-to-treat analysis, and 405 patients completed follow-up. Of the 405 patients completed follow-up, thirty patients were not included in the study due to the raw data hard disk error. Therefore, a total of 375 patients were included in the present study, with a mean age of 70.41±4.28 years and 52% of male (Table 1). There was no statistically significant difference between the patients included and excluded regarding the clinical characteristics and the group features (Supplement Table 1).

**Table 1.**
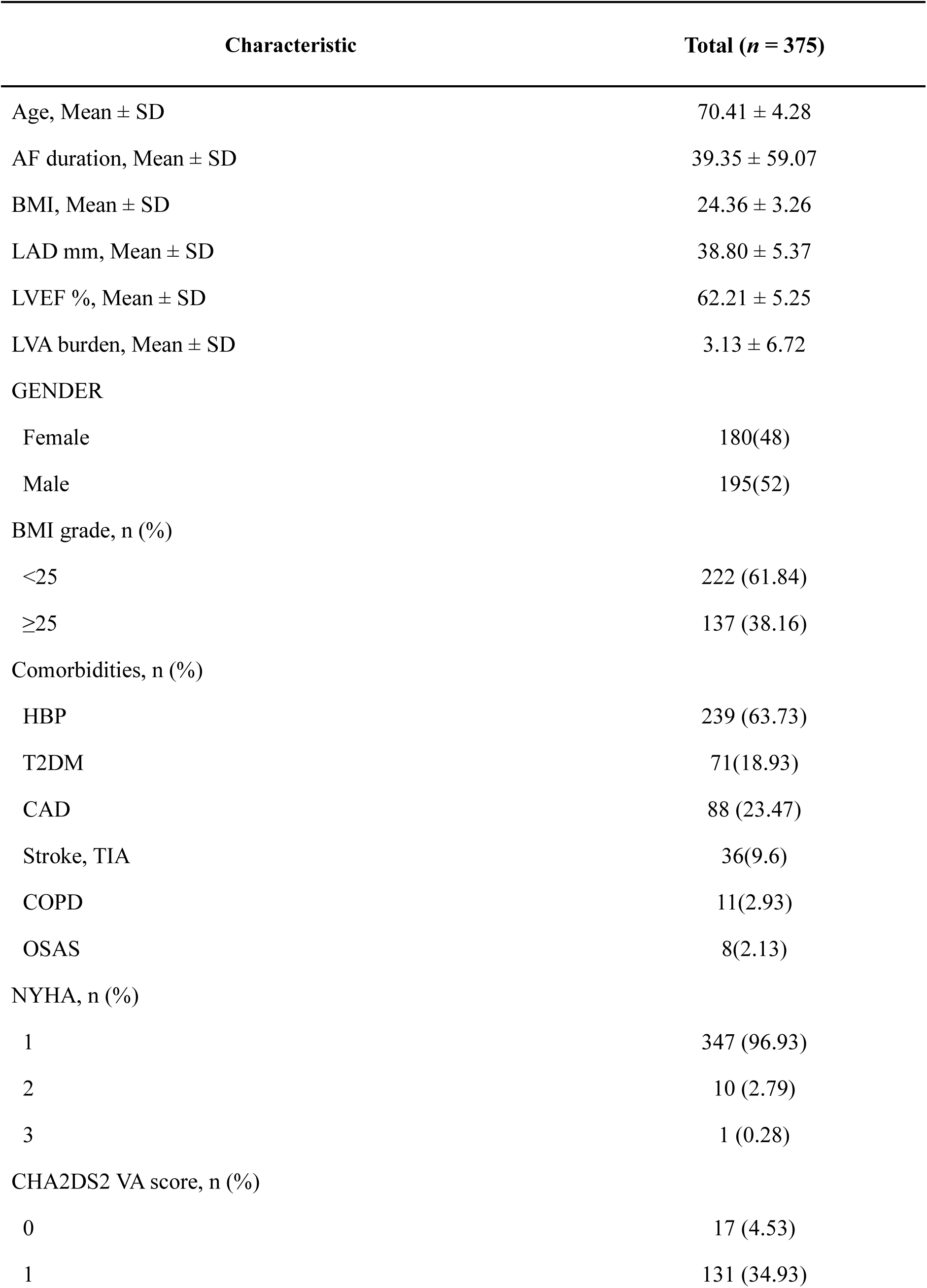
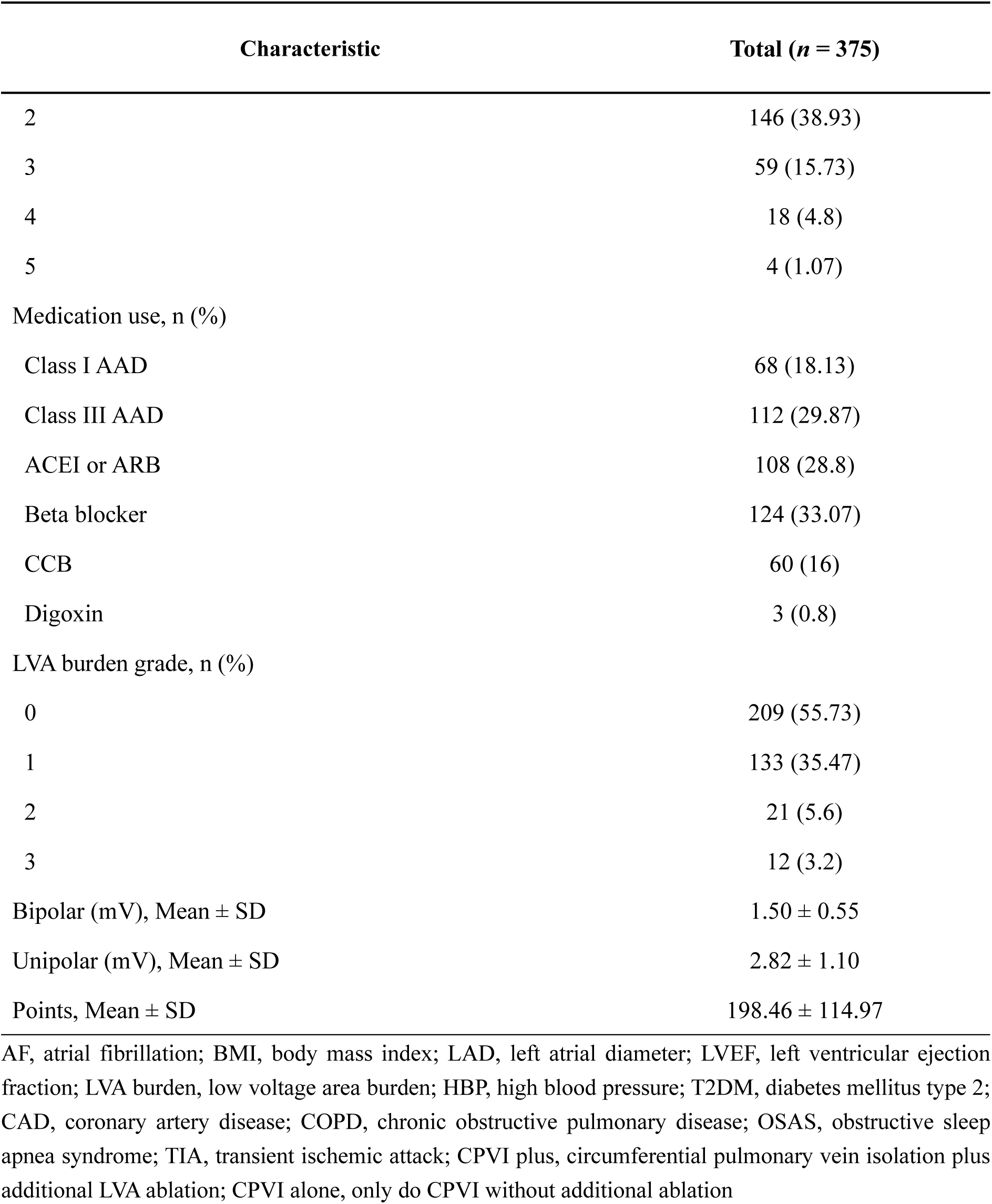
Baseline characteristics.

### Model development

The variables significantly associations with the long-term freedom from ATa were assessed by the univariate logistic regression and are shown in Table 2. Independent predictors were identified with use of stepwise multivariable analysis of logistic regression (Table 3). BV (OR= 0.18, 95%CI=0.10-0.33, P<0.001), a risk factor in univariable regression analysis, loses significance (BV in multivariate regression: OR=0.87, 95%CI=0.35-2.14, P=0.755) once UV (OR=0.12, 95% CI=0.06-0.25, P<0.001) is included in the multifactorial regression (Table 3). The adjusted predictive values for the recurrence of AF in relation to mean UV and BV are presented in Table 4. In the unadjusted model, UV (OR= 0.12, 95%CI=0.07-0.21, P<0.001) demonstrated better predictive recurrence effects and potential benefits when compared to BV (OR= 0.18, 95%CI=0.10-0.33, P<0.001). In the multivariable logistic regression model, adjustments were sequentially made for gender, age, left atrial dimension, CHA_2_DS_2_-VAS_C_ score and ablation strategy grouping. Even after these adjustments, the benefits persisted, with UV (OR= 0.11, 95%CI=0.06-0.20, P<0.001) consistently outperforming BV (OR= 0.22, 95%CI=0.09-0.31, P<0.001).

**Table 2.** Univariate logistic regression.

| Characteristics | $\beta$ | OR (95%CI) | *P* |
| --- | --- | --- | --- |
| Unipolar | -2.16 | 0.12 (0.07 - 0.21) | <.001 |
| Bipolar | -1.76 | 0.18 (0.10 - 0.33) | <.001 |
| LVA burden | 0.05 | 1.05 (1.02 - 1.09) | 0.003 |
| Group |  |  |  |
| CPVI alone |  | 1.00 (Reference) |  |
| CPVI plus | -0.70 | 0.49 (0.27 – 0.92) | 0.026 |

LAD, left atrial diameter; LVA burden, low voltage area burden; CPVI plus, circumferential pulmonary vein isolation plus additional LVA ablation; CPVI alone, only do CPVI without additional ablation; OR, odds ratio

**Table 3.** Multivariable logistic regression.

| Characteristics | $\beta$ | OR (95%CI) | *P* |
| --- | --- | --- | --- |
| Unipolar | -2.14 | 0.12 (0.06 - 0.25) | <.001 |
| Bipolar | -0.14 | 0.87 (0.35 - 2.14) | 0.755 |
| LVA burden | -0.00 | 1.00 (0.96 - 1.04) | 0.906 |
| Group |  |  |  |
| CVPI alone |  | 1.00 (Reference) |  |
| CPVI plus | -0.71 | 0.49 (0.26 – 0.92) | 0.026 |

**Table 4.**
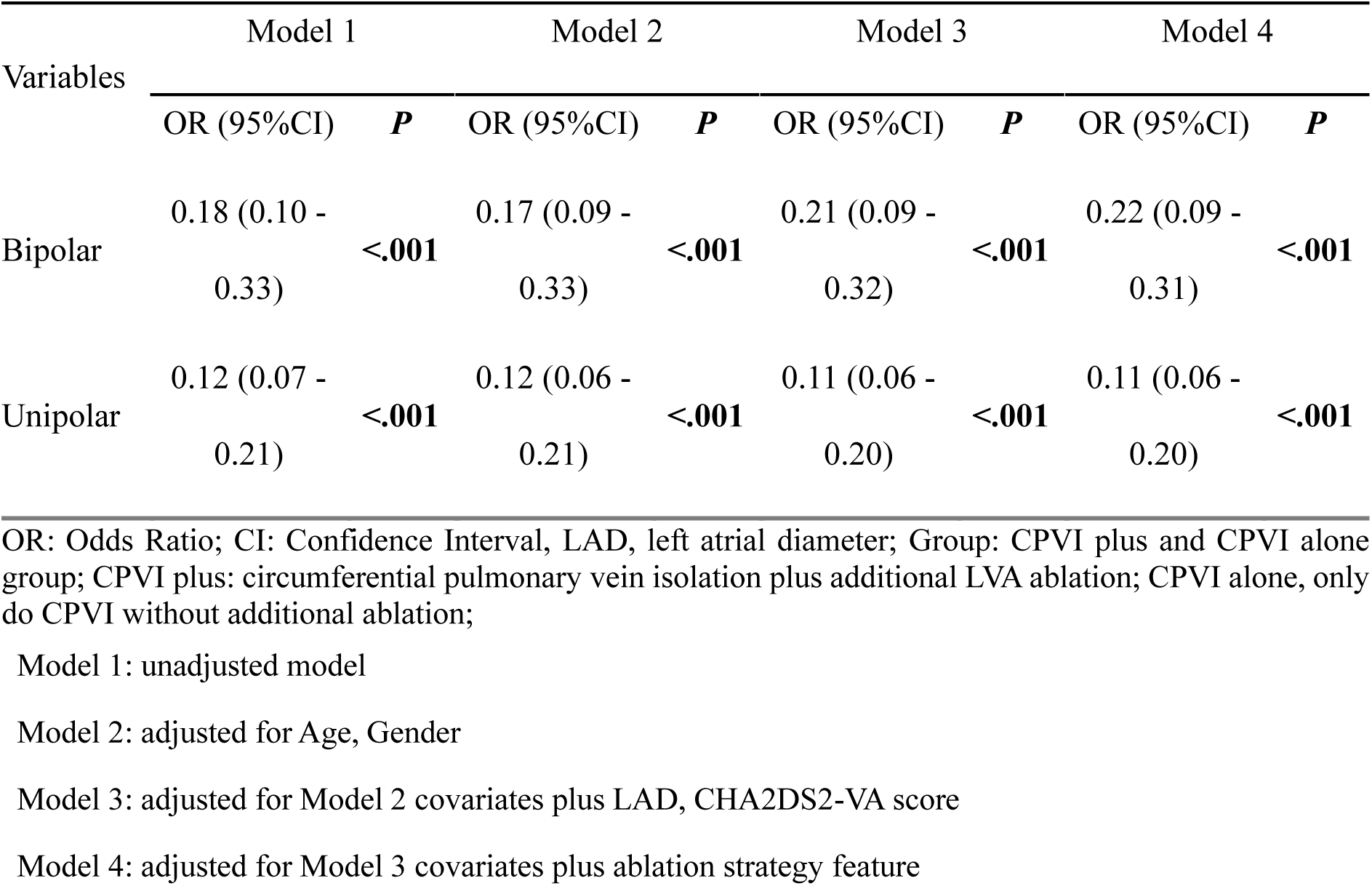
Freedom from atrial tachyarrhythmia recurrence of Bipolar voltage and Unipolar voltage adjusted for clinical features.

Multivariable logistic regressions were performed with UV and BV, respectively, and predictive models for UV and BV were established separately. UV (OR=0.11, 95%CI=0.06-0.19, p<0.001) (Table 5) and BV (OR=0.16, 95%CI=0.09-0.29, P<0.001) (Table 6) were discovered to be independently associated with AF recurrence and can establish their own model. The BV or UV from the CPVI alone/CPVI plus group was set as the independent variables to create two prediction models. The predict precision of the two models was then compared.

**Table 5.**
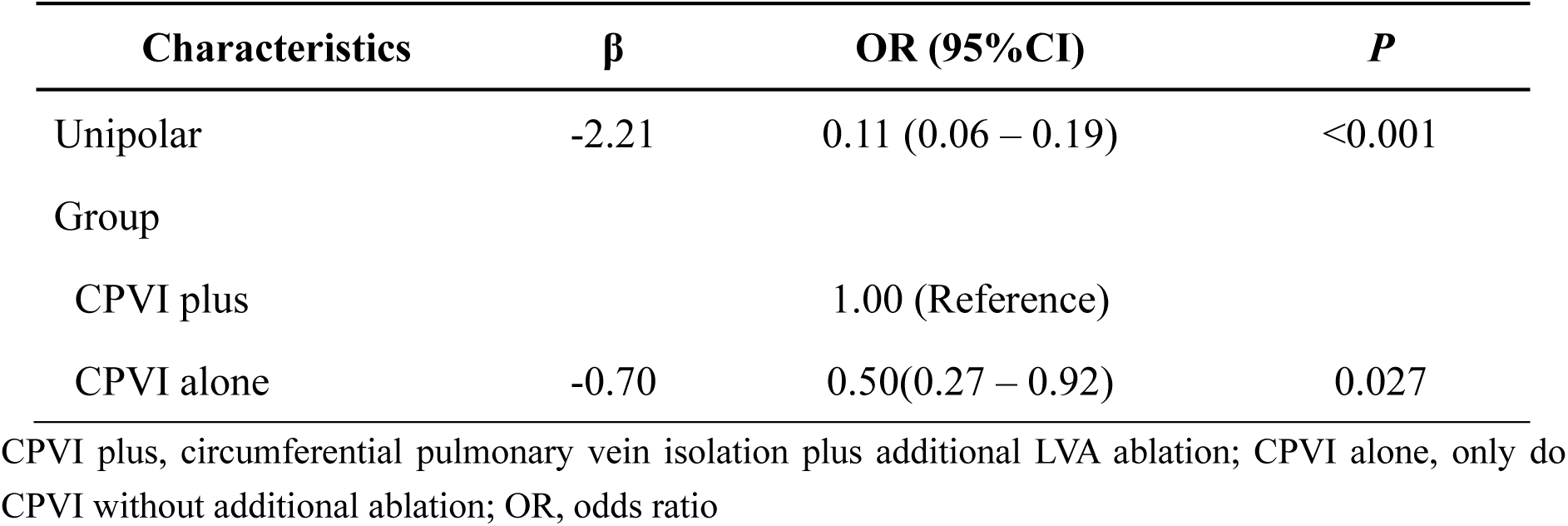
Multivariable logistic regression for Unipolar model.

**Table 6.** Multivariable logistic regression for Bipolar model.

| Characteristics | $\beta$ | OR (95%CI) | *P* |
| --- | --- | --- | --- |
| Bipolar | -1.84 | 0.16 (0.09 – 0.29) | <0.001 |
| Group |  |  |  |
| CPVI alone |  | 1.00 (Reference) |  |
| CPVI plus | -0.65 | 0.52 (0.30 – 0.92) | 0.023 |

The ROC analysis was used to investigate the discrimination of the prediction model. The AUC was 0.757 (95% CI:0.699-0.815) and 0.858 (95%CI:0.809-0.907) for BV and UV, and the difference in AUC values is statistically significant (P<0.001), respectively (Figure 1).

**Figure 1.**
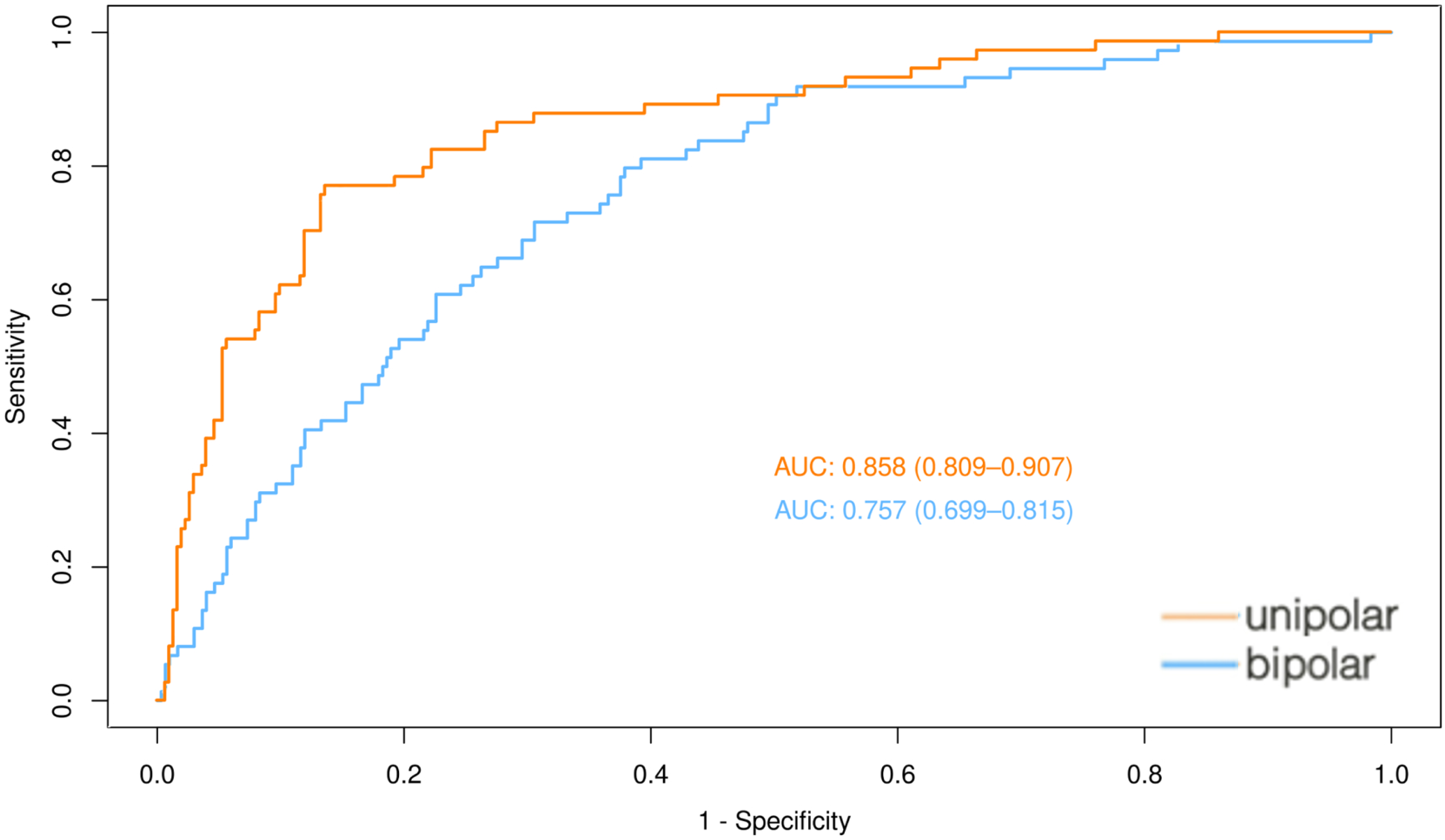
ROC for models in predicting recurrence in elder patients. ROC curve of bipolar and unipolar voltage models; The orange line shows the ROC for unipolar voltage, the blue line shows the ROC for bipolar voltage. AUC: area under the curve.

The calibration plots revealed good predictive accuracy between the actual probability and predicted probability of recurrence. The Hosmer and Lemeshow test showed an effective goodness-of-fit in both the BV model (P= 0.219, MAE=0.029) (Figure 2A) and the UV model (P= 0.3722, MAE=0.02) (Figure 2B).

**Figure 2.**
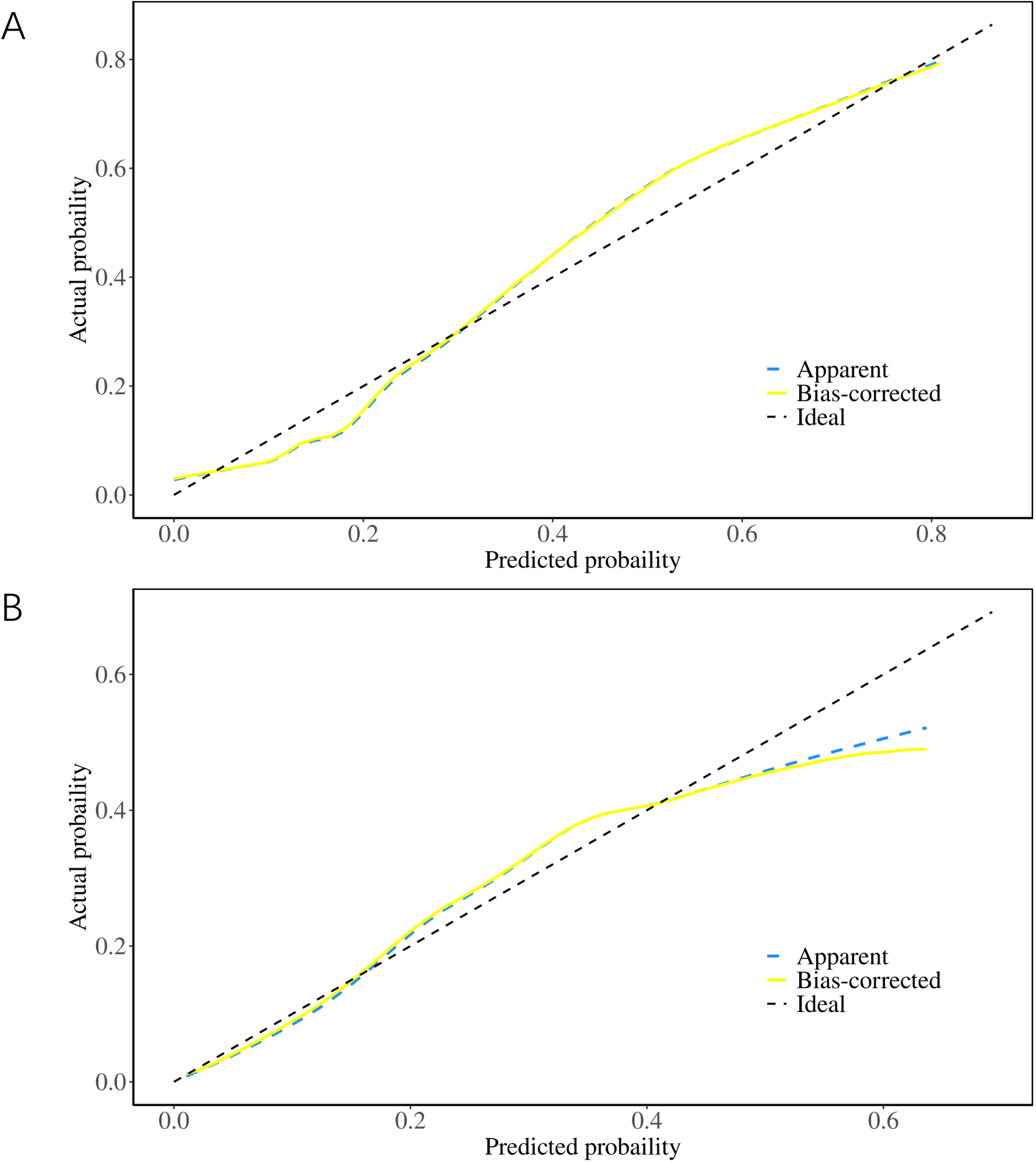
The Hosmer-–Lemeshow calibration curve for the model predicting probability of recurrence in elder AF patients. (A) Calibration curve for the bipolar voltage model. (B) Calibration curve for the unipolar voltage model. X-axis is predicted probability by model and y-axis is actual probability.

The decision curve analysis showed that the BV predicting model had a positive net benefit for a threshold probability between 0% and 45%. The UV predicting model has a better net benefit between 0% and 70% compared to BV predicting model (Figure 3).

**Figure 3.**
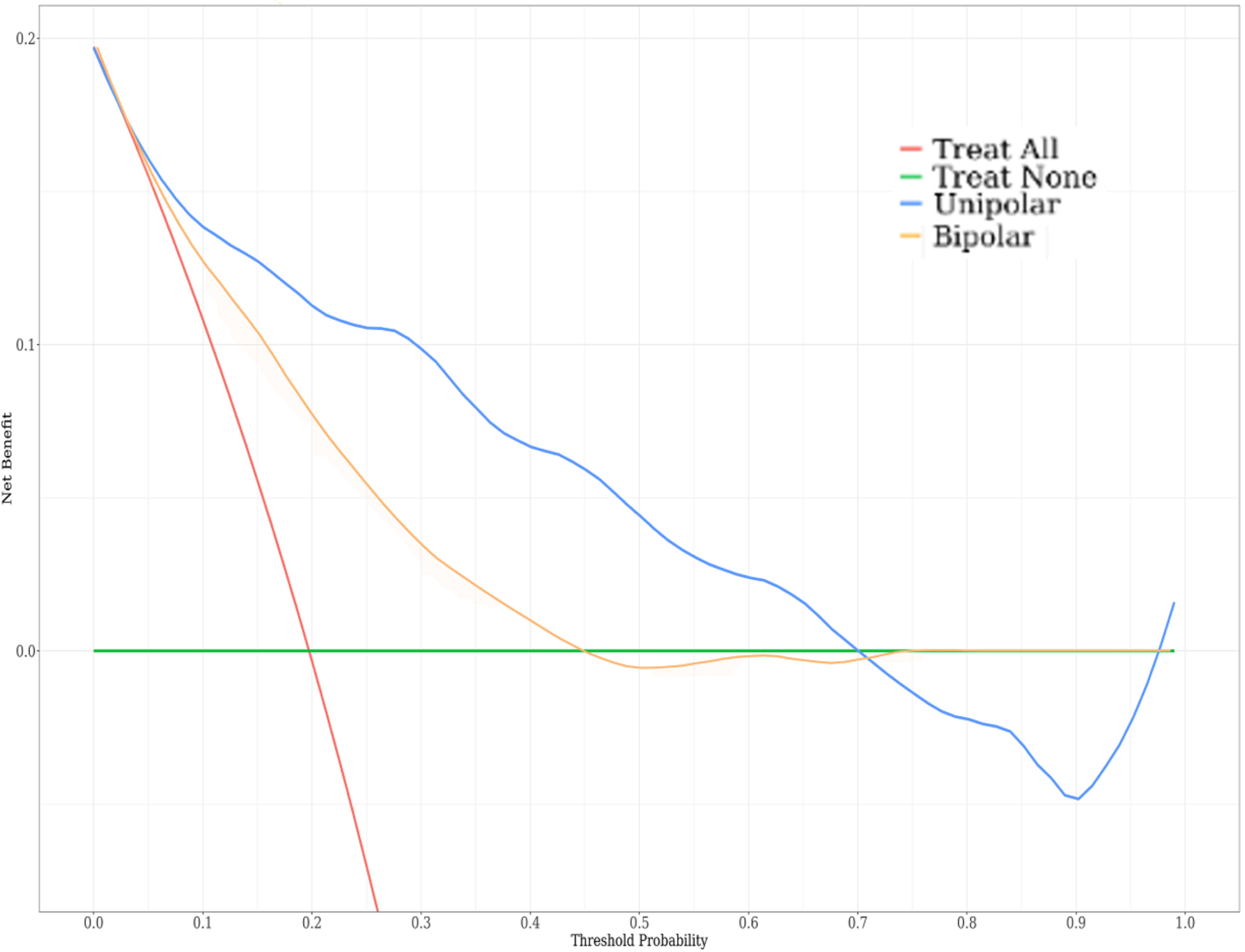
The decision curve analysis (DCA) for the model predicting probability of AF recurrence in elder AF patients. All the decision curve of the prediction models are composed of an X-axis which represents continuum of potential thresholds for risk and a Y-axis which represents the net benefit which is obtained by dividing the net true positives by the sample size. In the figure, the red curve “Treat ALL” shows the net benefit if all elder AF patients were intervened for bipolar or unipolar voltage. The green line “Treat None” shows the net benefit if no elder AF patients were intervened for bipolar or unipolar voltage. The blue and yellow line “prediction model for unipolar and bipolar voltage” curve shows the net benefit if it is used to select patients for intervention.

### Subgroup study

We then performed a subgroup analysis to explore the efficacy of predicting recurrence with BV and UV in the two subgroups of CPVI alone and CPVI plus (Supplement Table 2). The AUC was 0.751 (95% CI: 0.673-0.829) for BV and 0.843 (95% CI: 0.773-0.913) for UV in the CPVI alone cohort, and the difference between AUC values is statistically significant (P=0.0008), respectively (Figure 4A). In the meantime, the AUC was 0.750 (95% CI: 0.659– 0.841) for BV and 0.882 (95% CI: 0.809–0.954) for UV in CPVI plus cohort, and the difference between AUC values is statistically significant (P=0.0004), respectively (Figure 4B). The Hosmer and Lemeshow test of the UV model showed an effective goodness-of-fit both in CPVI alone cohorts(P=0.5471) (Figure 4C) and CPVI plus cohorts(P=0.3779) (Figure 4D). The Hosmer and Lemeshow test of BV model showed an effective goodness-of-fit both in CPVI alone cohorts(P=0.052) (Figure 4E) and CPVI plus cohorts(P=0.2132) (Figure 4F).

**Figure 4.**
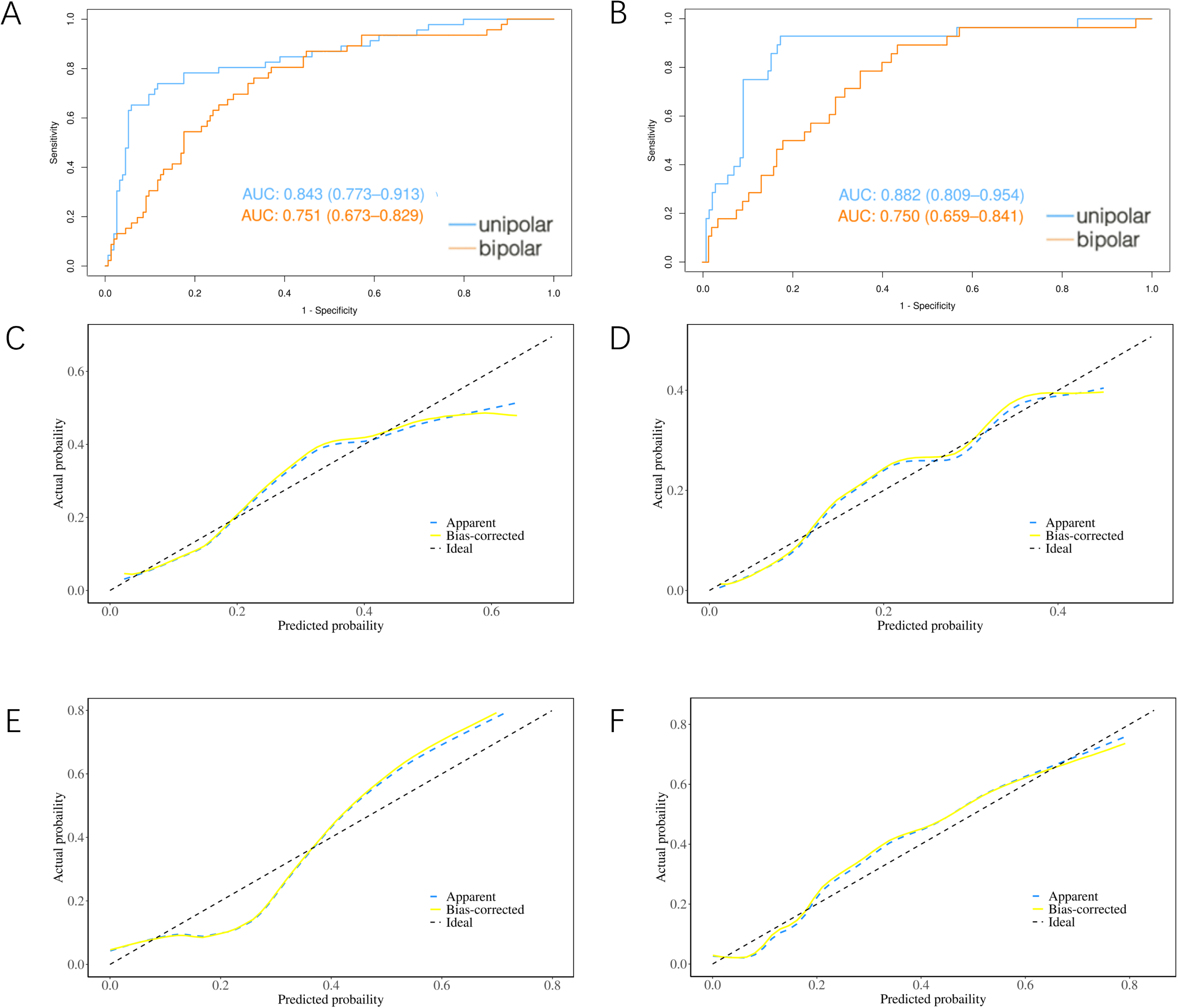
ROC for models in predicting AF recurrence in subgroup study. (A) ROC curve for the bipolar and unipolar voltage models in the CPVI alone group; (B) ROC curve for the bipolar and unipolar voltage models in the CPVI plus group. In both figures, the blue line shows the ROC for unipolar voltage, the yellow line shows the ROC for bipolar voltage. AUC: area under the curve. The Hosmer-–Lemeshow calibration curve for the model predicting probability of recurrence in elder AF patients. (C) Calibration curve for the unipolar voltage model in the CPVI alone group. (D) Calibration curve for the unipolar voltage model in the CPVI plus group. (E) Calibration curve for the bipolar voltage model in the CPVI alone group. (F) Calibration curve for the bipolar model in the CPVI plus group. X-axis is predicted probability by model and y-axis is actual probability of recurrence.

## Discussions

### Major finds

In this post-hoc analysis of the STABLE-SR III trial, we investigated and compared the predictive efficacy of the mean LA UV vs BV for recurrent ATa after AF ablation in elderly paroxysmal AF patients. Our main findings include: (1) Both the mean unipolar and bipolar left atrial voltage serve as risk indicators for ATa events after AF ablation. However, only the mean UV is independently associated with the long-term freedom from ATa. (2) The predictive value of UV for recurrence is significantly superior to that of BV, as demonstrated by the model developed through multivariable analysis. These results underscore the advantages of unipolar mapping in characterizing the comprehensive three-dimensional substrate pattern.

### Role of unipolar vs bipolar voltage in detecting atrial fibrosis

Myocardial fibrosis plays a crucial role in the cardiac remodeling observed in individuals with AF^2^. Cardiac fibroblasts transmit electrical currents between myocardial cells through connecting proteins, resulting in uneven current conduction, slowed conduction velocity, shortened action potential and increased conduction heterogeneity -- all of which can be reflected in the intracardiac electrograms obtained through contact mapping^12–16^. Deneke T et al. were the first to demonstrate a histopathological correlation of BV mapping in identifying the ventricular arrhythmic substrate^17^. Although several studies have highlighted the utility of bipolar recording in detecting endocardial and epicardial scars in both ischemic and non-ischemic cardiomyopathy, inconsistency between BV and the transmural extent of infraction has been reported. This discrepancy calls into question the “field of view” of bipolar electrograms^9, 10, 18^. An elegant study introduced UV mapping and demonstrated its superiority in characterizing transmural substrate features in non-ischemic cardiomyopathy^9^.

The human atrial wall is a thin structure, with average measurements of wall thickness between 1 and 4 mm but with a broader range extending from 0.5 to 12 mm^19^. Furthermore, several studies have provided evidence for endocardial-epicardial voltage and activation asynchrony in patients with AF^8, 18, 20, 21^. BV, with its detection range limited to the endocardium or sub-endocardium -- sometimes within 1-2 mm deep layers, might not be optimal for capturing the complete substrate features of the entire layer. Consequently, UV mapping might also hold an advantage in reflecting three-dimensional substrate information in atrium. In the present study, for the first time, the efficacy of UV in predicting the recurrence of ATa after catheter ablation was demonstrated to be superior to that of BV. Admittedly, in univariable analysis, both UV and BV correlated with the long-term freedom of ATa. However, in multivariable analysis, the association between BV and outcome was overshadowed by the stronger link between UV and outcome. Furthermore, the model developed based on the UV demonstrated a larger area under the ROC for predicting the ATa recurrence compared to BV. Theoretically, this statistical superiority can only be attributed to the previously mentioned the wider “field of view” of UV for deeper substrates.

It’s worth noting that the superiority of UV over BV for predicting recurrence exists regardless of whether patients receiving substrate modification, which is in line with the findings in DECAFF study. In DECAFF-II trial, atrial fibrosis detected by MRI, which is also a comprehensive approach, is similar to our UV assessment and shares the analogues predictive value in predicting the AF recurrence.

### Clinical implications

To quantify atrial fibrosis in AF patients who underwent ablation is of clinical importance. Firstly, it aids in identifying those patients who were most likely to benefit from the procedure. In DECAAF studies, severer atrial fibrosis has been proven to be a significant predictor for ablation failure. Cardiac magnetic resonance imaging (CMRI) showed potential utility in evaluating atrial fibrosis^14, 16^. However, this technique has not been widely adopted, and CMRI is time-consuming and not readily available. BV during the ablation procedure is often adopted as a simple and convenient tool to predict the long-term outcomes. Herein, we have developed a model integrating the unipolar and clinical parameters as a more powerful tool compared to BV. Whether the plus LVA modification was performed or not, the mean left atrium UV of 2.82±1.10mV provided a C-index of 0.858 (95%CI:0.809-0.907). Another potential application scenario of the UV mapping for AF ablation is the patient-tailored ablation approach based on the LVA. To date, there is inconsistency regarding the benefits of fibrosis-targeting ablation approach^7, 16, 22^. The DECAFF study demonstrated no significant difference in terms of ATa recurrence between the CMRI-guided fibrosis ablation plus PVI and PVI only^14, 16^. However, other studies, including STABLE-SR-III, have shown positive results^7, 22^. The discrepancy in fibrosis identification methods may contribute to this inconsistency. We have long observed a discordance between CMRI-based fibrosis assessment and that of LVA assessed by bipolar recording. In the DECAFF series, nearly half of the patients were categorized as stage 3 or stage 4 (at least 20% of the atrial wall has fibrosis), while in STABLE-SR series, only about 10% patients had >10% of the atrial wall identified as confluent LVA^5–7, 14, 16^. We speculate that the limited “field of view” by bipolar mapping partly contributes to this discordance. In the future, it seems reasonable and promising to employ the UV-based LVA as the surrogate for fibrosis during this patient-tailored ablation strategy.

### Limitations

As a post-hoc analysis of the STABLE-SR-III research, is inevitably constrained by the inclusion criteria, specifically focusing on data from elderly patients with paroxysmal AF. Consequently, the findings cannot be readily extrapolated to the entire population. Furthermore, the study did not explore the determination of precise UV cutoff value to standardize the low-voltage area and discuss its relationship with prognosis.

## Conclusion

The endocardial mean UV of LA demonstrates a higher predictive value for predicting recurrence after ablation compared to mean BV in elderly patients with AF. The superiority of unipolar mapping in predicting recurrence implicates its advantage of providing a broader, more penetrative field of view to identify arrhythmogenic substrates in deeper layers of the atrium.

## Data Availability

The data that support the findings of this study are available from the corresponding author, [Weizhu Ju], upon reasonable request.

## Non-standard Abbreviations and Acronyms

AF: atrial fibrillation
ATa: atrial tachyarrhythmia
BV: bipolar voltage
CI: confidence interval
CPVI: circumferential pulmonary vein isolation
OR: odds ratio
LA: left atrium
LAD: left atrial diameter
LVA: low-voltage area
UV: unipolar voltage

## Acknowledgement

We thank all patients for participating in this study and all clinicians for their invaluable assistance in the evaluation of mapping data and clinical assessments.

## Supplement material

Table S1, S2

---

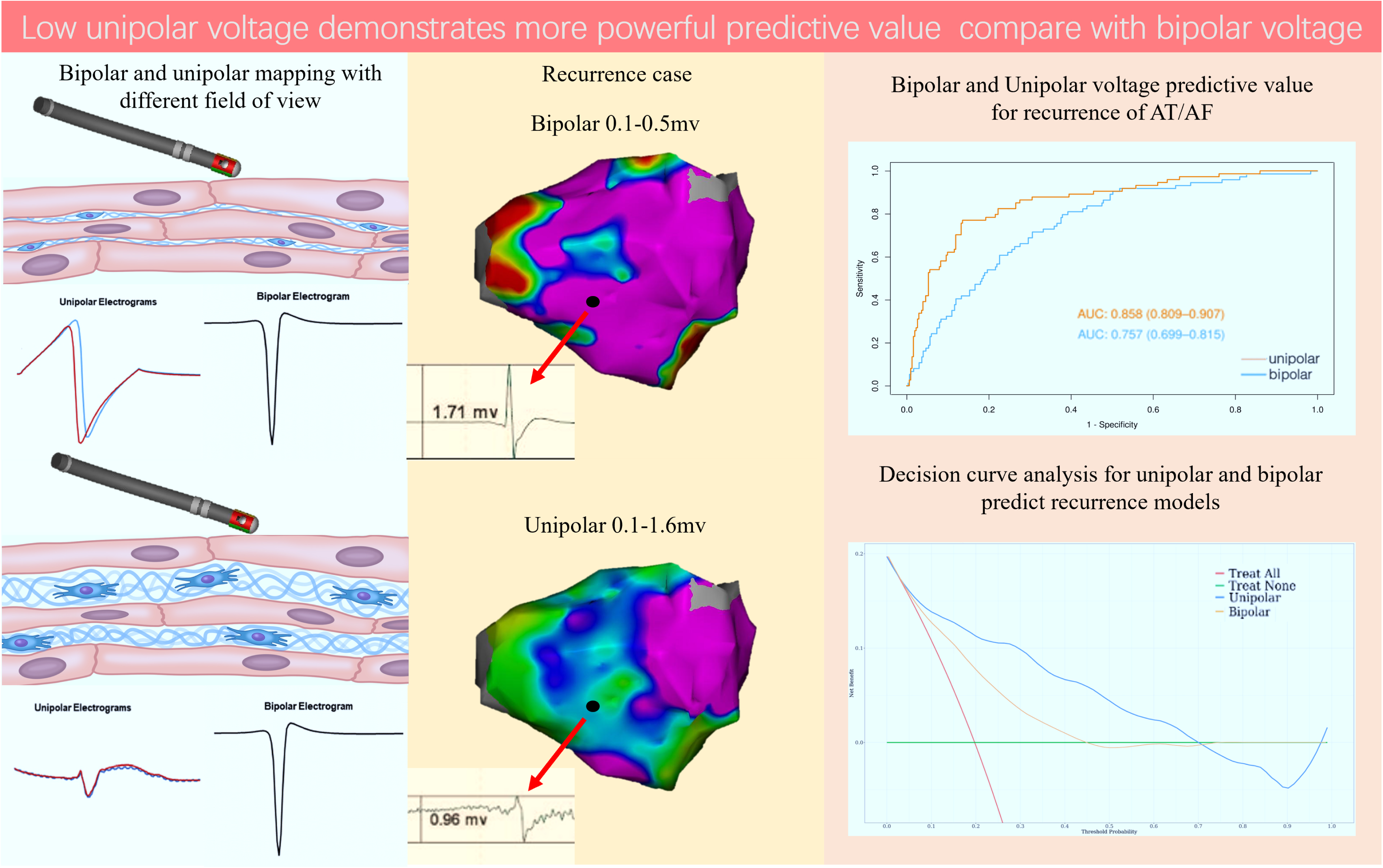

